# MRI-based cortical gray/white matter contrast in young adults who endorse psychotic experiences or are at genetic risk for psychosis

**DOI:** 10.1101/2024.05.28.24308025

**Authors:** Nasimeh Naseri, Dani Beck, Lia Ferschmann, Eira R. Aksnes, Alexandra Havdahl, Maria Jalbrzikowski, Linn B. Norbom, Christian K. Tamnes

## Abstract

Research has reported group-level differences in cortical grey/white matter contrast (GWC) in individuals with psychotic disorders. However, no studies to date have explored GWC in individuals at elevated risk for psychosis. In this study, we examined brain microstructure differences between young adults with psychotic-like experiences or a high genetic risk for psychosis and unaffected individuals. Moreover, we investigated the association between GWC and the number of and experiences of psychosis-like symptoms. The sample was obtained from the Avon Longitudinal Study of Parents and Children (ALSPAC): the psychotic experiences study, consisting of young adults with psychotic-like symptoms (n = 119) and unaffected individuals (n = 117), and the schizophrenia recall-by-genotype study, consisting of individuals with a high genetic risk for psychosis (n = 95) and those with low genetic risk for psychosis (n = 95). Statistical analyses were performed using FSL’s Permutation Analysis of Linear Models (PALM), controlling for age and sex. The results showed no statistically significant differences in GWC between any of the groups and no significant associations between GWC and the number and experiences of psychosis-like symptoms. In conclusion, the results indicate there are no differences in GWC in individuals with high, low or no risk for psychosis.

## 1. Introduction

Magnetic resonance imaging (MRI) studies have documented group-level brain structural and microstructural differences between individuals with psychotic disorders and unaffected individuals (Birur et al. 2017; Bose et al. 2009; Ching et al. 2022; Falkai, Schmitt, and Andreasen 2018; Schmidt et al. 2016; Andreassen et al. 2023; Haukvik et al. 2018; Hibar et al. 2018). However, whether these differences represent risk factors or develop together with the disorder or in relation to medication is not well understood. Studies of individuals at risk, whether it be a genetic risk or individuals who endorse subclinical psychosis symptoms, can provide new insights into developmental processes in psychotic disorders. In the present study, we utilized a state-of-the-art MRI-based signal intensity measure, gray/white matter contrast (GWC), and explored brain microstructure in young adults with genetic high risk or those endorsing psychotic experiences in comparison to unaffected individuals. Additionally, we tested whether individual differences in GWC in these risk groups were related to the number and experiences of psychosis-like symptoms.

Patients with schizophrenia and bipolar disorder have been found to have widespread structural brain differences compared to unaffected individuals. Both groups, on average, show lower cortical volume, with a greater reduction in volume in schizophrenia affecting frontal, temporal and occipital regions (Madre et al. 2020; Rimol et al. 2012). Patients with schizophrenia also show widespread thinner cortices and smaller cortical surface area, with the largest effects observed in the frontal and temporal regions (van Erp et al. 2016). In patients with bipolar disorder, studies predominantly result thinner cortices, but not smaller surface area, in comparison to unaffected individuals (Hibar et al. 2018; Madre et al. 2020). Patient groups also show, on average, larger ventricles, and smaller bilateral hippocampi and left thalamus (Haukvik et al. 2018; Hibar et al. 2018; van Erp et al. 2016), though patients with schizophrenia appear to have smaller hippocampal and amygdala volumes and larger right putamen volume in comparison to patients with bipolar disorder (Haukvik et al. 2018; Ohi et al. 2022; van Erp et al. 2016).

One approach to determine whether the observed differences in brain structure in patients with psychotic disorders precede illness onset is to study individuals at elevated risk for psychosis (Paolo Fusar-Poli 2017). This is particularly important for understanding the pathological mechanisms driving clinical and functional deterioration commonly seen in psychosis, which could inform the development of interventions aimed at preventing progression from a high-risk state to a clinical disorder (Cannon 2008). Individuals can be considered at high genetic risk for developing a psychotic disorder based on familiar risk (Smieskova et al. 2010a) or the presence of common or rare genetic variants associated with psychotic disorders (Karayiorgou, Simon, and Gogos 2010). Individuals from the general population who endorse psychotic experiences (PE), as well as help-seeking individuals who do not meet diagnostic criteria for a psychotic disorder, are also considered at increased risk for developing psychosis (Paolo Fusar-Poli 2017; Modenato et al. 2021; Yung et al. 1996; Jalbrzikowski et al. 2021; Drakesmith et al. 2015).

Monozygotic twins of individuals with a psychotic disorder are estimated to have a 40-50% lifetime risk of developing a psychotic disorder, while first-degree relatives of patients with schizophrenia have an approximately 10-fold increased risk for later illness (Smieskova et al. 2010). Familial risk for schizophrenia has been associated with smaller brain volumes and lower cortical thickness relative to subjects from low-risk families (R. C. K. Chan et al. 2011; Ivleva et al. 2013; Zwarte et al. 2019). Genetic risk can also be studied using polygenic risk scores (PRS). The PRS is a result of a large number of common genetic variants that are likely contributing to a disorder (Ripke et al. 2014). Each variant has a small effect, but by combining them one can derive a score with larger predictive power (Choi, Mak, and O’Reilly 2018). These PRSs can be calculated in unaffected individuals to study the generic risk of various diseases (Daetwyler, Villanueva, and Woolliams 2008). High PRS for schizophrenia has – in unaffected adults – been associated with lower cortical thickness in lateral orbitofrontal, inferior frontal, and posterior cingulate regions (Zhu et al. 2021). Contrary to these findings, however, a meta-analysis (van der Merwe et al. 2019) did not find any significant associations between PRS for schizophrenia and grey matter volume, white matter volume, globus pallidus volume and total brain volume.

The risk of developing a psychotic disorder ranges from 41% to 54% within one year and 18% to 20% within two years among individuals with clinical high risk for psychosis (CHR) (Paolo Fusar-Poli 2017; Paolo Fusar-Poli et al. 2015; 2020; Jalbrzikowski et al. 2021). A large body of work has used MRI to investigate brain structural differences in individuals at CHR for psychosis (Chung et al. 2019; Del Re et al. 2021; Fornito et al. 2008; P. Fusar-Poli et al. 2011; Iwashiro et al. 2012; Klauser et al. 2015; Koutsouleris et al. 2009; Kwak et al. 2019; Mechelli et al. 2011; Sun et al. 2009; Takayanagi et al. 2017; Tomyshev et al. 2019; Velakoulis et al. 2006; Ziermans et al. 2009; Zikidi et al. 2020). The largest to-date study of CHR individuals found that in comparison to unaffected individuals, individuals with CHR showed widespread lower cortical thickness, but no differences in surface area or subcortical volumes (Jalbrzikowski et al. 2021). Moreover, CHR individuals who later developed a psychotic disorder had lower cortical thickness in paracentral, superior temporal and fusiform regions in comparison to both CHR individuals who did not develop psychosis and unaffected individuals (Jalbrzikowski et al. 2021). Similar structural differences have been found among young adults who endorse psychotic experiences compared to unaffected individuals. For example, studies have found group-level differences in brain microstructures such as fractional anisotropy (Peters and Karlsgodt 2015; Smigielski et al. 2022; León-Ortiz et al. 2022; Drakesmith et al. 2016), differences in grey matter volume in the left supramarginal gyrus (Drakesmith et al. 2015), and greater brain volume in the middle frontal gyrus into the superior frontal gyrus (Fonville et al. 2019). However, another study found no differences in gray matter volume in a whole-brain analysis (Fonville et al. 2015).

Although the studies discussed above indicate that differences in brain structure exist prior to the onset of psychotic disorders, newer and less explored MRI signal intensity measures can provide additional information about brain microstructure (Norbom et al. 2019). These might offer increased biological specificity, as GWC is presumed to be a proxy measure for differences in intracortical and subjacent white matter myelin content (Eickhoff et al. 2005; Salat et al. 2009; Stüber et al. 2014). One such approach is to compute GWC from intensities sampled across the cortical mantle and within closely subjacent white matter (Salat et al. 2009), where higher GWC indicates a greater discrepancy between grey and white matter. Examining GWC among young adolescents could improve the sensitivity to detect differences between individuals at risk for psychosis and unaffected individuals. This period aligns with the protracted maturation of subjacent myelination, estimated to continue until the late 20s or early 30s (Drakulich et al. 2021; Norbom et al. 2020; Westlye et al. 2010). This period of myelin maturation also aligns with the vulnerability period for psychotic disorders, and dysmyelination has been proposed as one possible mechanism for these disorders (Bartzokis 2003; Whitford et al. 2012). Abnormalities in myelination have been associated with dysfunctional connectivity between the frontal lobes and extra frontal and subcortical structures in schizophrenia (Mighdoll et al. 2015).

During childhood and adolescence, GWC shows an age-related decrease, thought to partly reflect protracted intracortical myelination (Norbom et al. 2019). Only a few studies to date have used GWC to examine brain microstructure in patients with psychotic disorders (Jørgensen et al. 2016; Kong et al. 2012; 2015; Chwa et al. 2020), while no studies have examined GWC in psychosis risk groups. A study by Jørgensen et al. (2016) including adults with schizophrenia, bipolar disorder and unaffected individuals found higher GWC in pre- and postcentral gyri, the transverse temporal gyri, posterior insula, and parieto-occipital regions in patients with schizophrenia. The study also found higher contrast primarily in the left precentral gyrus in patients with bipolar disorder, but no significant differences between the two patient groups. Moreover, Jørgensen et al. (2016) found that increased GWC was associated with increased severity of hallucinations in patients, but no associations for delusions or medication. A study by Chwa et al. (2020) similarly showed, that in comparison to unaffected individuals, individuals with schizophrenia had higher GWC in the right superior frontal lobe encompassing the sensorimotor region. Additionally, they observed an association between lower GWC and increased exposure to second-generation antipsychotics within the superior frontal lobes. A large population-based study of youth by Norbom et al. (2019) also found that increased GWC was associated with more psychosis-spectrum symptoms. Contrary to the results from these three studies, Kong et al. (2012) compared GWC in a small sample of adult patients with schizophrenia and unaffected individuals and observed lower GWC in the patient group in large portions of the cortex, including frontal, temporo-parietal and lateral occipital regions. In a follow-up study, Kong et al. (2015) also found lower regional GWC in the patient group. In sum, the three largest existing studies indicate increased GWC primarily in highly myelinated sensory and motor regions in patients with schizophrenia and increased GWC to be associated with psychosis symptoms, although discrepant findings have also been reported. Our understanding of the progressive effect these disorders have on GWC is however limited as no studies to date have examined GWC in individuals with genetic risk for psychosis or those who endorse psychotic experiences.

The present study aimed to explore differences in brain microstructure before the onset of psychotic disorders by comparing GWC between young adults with genetic risk, those endorsing psychotic experiences, and unaffected individuals. We hypothesized that we would observe regionally higher GWC in the psychotic experience group compared to unaffected individuals, based on previous findings (Chwa et al. 2020; Jørgensen et al. 2016; Norbom et al. 2019). To our knowledge, there is a lack of research on GWC in groups with genetic risk for psychosis, with previous studies reporting limited evidence for associations between PGS for schizophrenia and brain macrostructure (Lancaster et al. 2019; Papiol et al. 2014; van der Merwe et al. 2019). Furthermore, no association between risk for schizophrenia and genes associated with myelination have been found (Goudriaan et al. 2014; Stokowy et al. 2018).

Thus, we hypothesized that there would be no significant difference in GWC between the high genetic risk group and the low genetic risk group. We also hypothesized regionally higher GWC in the group of individuals who endorsed psychotic experiences relative to the genetic risk group. Finally, we hypothesized that individual differences in the number and experiences of psychosis-like symptoms would be positively associated with the GWC in the psychotic experiences group, based on findings from a previous study observing a positive association between hallucination symptoms and GWC in patients with psychotic disorders (Jørgensen et al. 2016). We did not expect to observe a similar association in the genetic risk group since previous studies have reported no association between PRS for schizophrenia and psychotic symptoms (Jones et al. 2016; Lancaster et al. 2019). Overall, the current study aimed to provide new knowledge of the neural mechanisms underlying risk for psychotic disorders and investigate whether previously reported differences in GWC in patients represent pre-existing risk factors or emerge later in the course of the disorders.

## 2. Experimental procedures

### 2.1 Sample

The data for this study is drawn from the Avon Longitudinal Study of Parents and Children (ALSPAC) study at the University of Bristol (see Golding, Pembrey, and Jones (2001) for more information). Pregnant women residents in Avon, UK with an expected date of delivery between the 1^st^ April 1991 and the 31st December 1992 were invited to take part in the ALSPAC study. The initial number of pregnancies enrolled was 14,541 and 13,988 children were alive at 1 year of age. When the oldest children were approximately 7 years of age, an attempt was made to bolster the initial sample with eligible cases who had failed to join the original study. The total sample size for analysis using any data collected after the age of seven is therefore 15,447 pregnancies and 14,901 children alive at 1 year of age. Data of 14,822 unique women (Generation 0 mothers) were enrolled in ALSPAC as of September 2021 (Boyd et al. 2013; Fraser et al. 2013).

The final sample in the current study included participants from two ALSPAC MRI sub-studies, the Psychotic Experiences (PE) study and the schizophrenia recall-by-genotype (SZP-RbG) study (Sharp et al., 2020) and consisted of 119 (83 female, 36 male) individuals with psychotic-like experiences, 117 (73 female, 44 male) unaffected individuals, 95 (50 female, 45 male) individuals with a high genetic risk for schizophrenia, and 95 (50 female, 45 male) with a low genetic risk for schizophrenia.

The PE study (Drakesmith et al. 2016) included individuals with previous psychotic-like experiences, defined as having experienced at least one definitive or suspected psychotic experience during the past six months at the age of 17-18 years, assessed using the psychotic-like symptoms semi-structured interview (PLIKS), along with unaffected individuals. A total of 252 participants were recruited for MRI scanning between the ages of 21 and 24 years.

The SZP-RbG study included individuals with high or low genetic risk for schizophrenia, defined by having high or low PRS for schizophrenia. Genotyping was done by subcontracting the Wellcome Trust Sanger Institute, Hinxton, U.K., and the Laboratory Corporation of America, Burlington, North Carolina, U.S. The genetic risk scores for schizophrenia were calculated using methods described by the International Schizophrenia Consortium, based on the older version of the Psychiatric Genomics Consortium summary statistics (PGC) SCZ genome-wide association studies (GWAS) (Ripke et al. 2014). The genetic risk scores for schizophrenia were derived from Plink (version 1.07; as described elsewhere (Lancaster et al. 2019), which summed the number of risk alleles for each single nucleotide polymorphism (SNP). In total, 196 individuals were recruited for MRI scanning between the ages of 19 and 21 years. Researchers were blinded to which tail of the genetic risk score for schizophrenia distribution participants were selected from during both data collection and processing.

Before analyses, we excluded duplicate datasets from the PE subsample (n = 6) and participants with missing PLIKS data at age 24 (n = 10). Further details on the final samples after these exclusions and exclusions based on MRI quality control as described below (n = 6) (Section 2.4) are provided in **Table 1**.

**Table 1.**
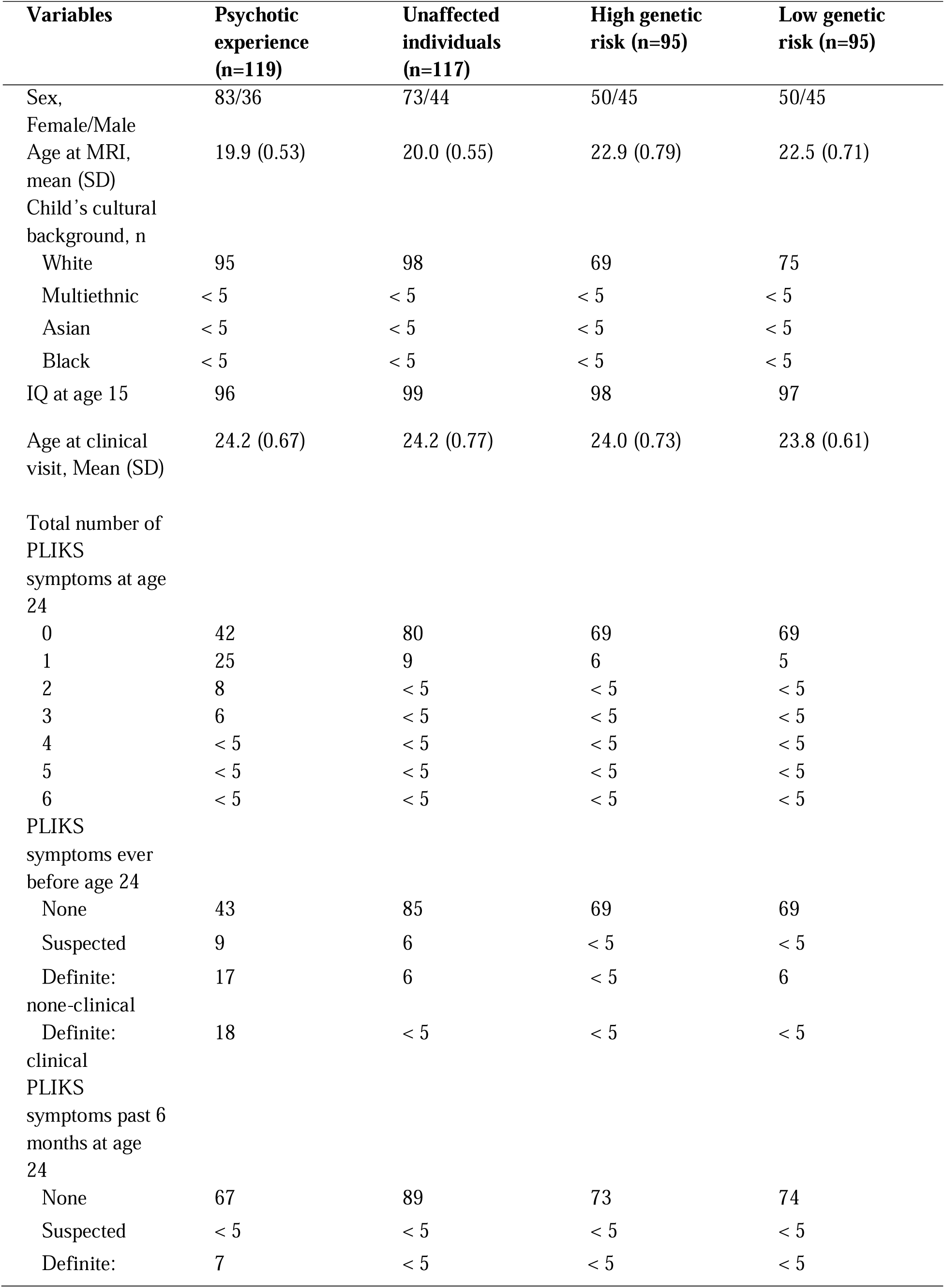
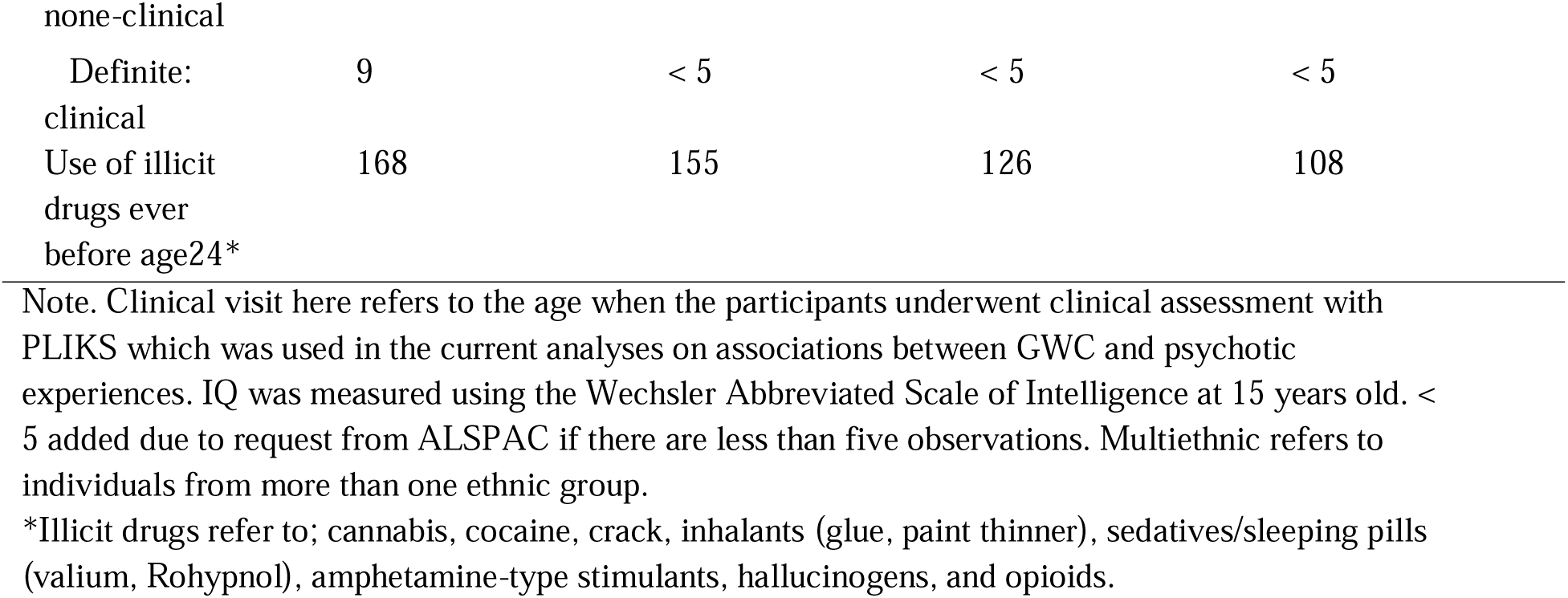
Sample demographics.

Written informed consent was collected from all participants. Ethical approval for all neuroimaging studies was obtained from the ALSPAC Ethics and Law Committee and the Local Research Ethics Committees. Consent for biological samples has been collected in accordance with the Human Tissue Act (2004). The current study was conducted in line with the Declaration of Helsinki and was approved by the Norwegian Regional Committee for Medical and Health Research Ethics (REK 269241).

### 2.2 MRI acquisition

All the neuroimaging data were acquired at Cardiff University Brain Research Imaging Center (CUBRIC) on the same 3 Tesla General Electric HDx scanner (GE Medical Systems) using an 8-channel head coil. The T1-weighted structural images in both MRI sub-studies were obtained using an FSPGR sequence (1 mm isotropic resolution, TR = 7.8/7.9 ms, TE = 3.0 ms, TI = 450 ms, flip angle = 20°). Participants were instructed to have a typical night’s sleep before each scan, not to drink more than one alcoholic beverage and to abstain from drinking coffee within 2 hours preceding the scan (Sharp et al. 2020).

### 2.3 MRI processing

FreeSurfer (version 7.3.2) was used to process the T1-weighted images via the ‘recon-all’ command including the -qcache flag. This automated pipeline includes removal of non-brain tissue, voxel intensity correction for B1 field inhomogeneities, segmentation of voxels into white matter, grey matter or cerebral spinal fluid and generation of surface-based models of white and grey matter (Fischl 2012).

We sampled signal intensities for each participant from the nonuniform intensity normalized volume (nu.mgz) using the FreeSurfer function mri_vol2surf. For each vertex, gray matter intensities were sampled at six equally spaced points. White matter intensities were sampled at each vertex at 10 equally spaced points, starting from the gray/white boundary and ending at a fixed distance of 1.5 mm into the white matter. Using an established procedure (Jørgensen et al., 2016), we calculated the average intensity value for each tissue type to obtain single separate measures of gray and white matter intensity per vertex. GWC was then computed as 100X (white-gray)/[(white+gray)/2], such that a higher value reflects a greater difference between cortical gray matter and white matter signal intensities.

### 2.4 MRI quality control

We performed a manual rating of reconstructed images following the ENIGMA consortium protocol (https://enigma.ini.usc.edu/protocols/imaging-protocols/). Visual inspection of the cortical external parcellation and cortical internal parcellation was performed separately for all the participants. Here, images were rated a “pass”, “moderate”, or “fail” quality, where fail was defined as the presence of motion or other artefacts that significantly compromised image quality. For our analyses, focusing on GWC, we used the cortical quality control ratings.

Three subjects in the PE sub-study and three subjects in the SCZ-RbG sub-study failed the quality control, and these six participants were subsequently excluded from all analyses.

### 2.5 Clinical assessment

Psychotic experiences in the high-risk groups of both sub-studies (the Pe study and the SZP-RbG study) were assessed using PLIKS, conducted around the age of 24 years (mean = 24.2, SD = 0.7). PLIKS is a semi-structured interview which consists of 12 core questions covering the past 6-month occurrence of hallucinations, delusions, and experience of thought interference. Seven of the 12 core questions were derived from the Diagnostic Interview Schedule for Children-IV (DISC-IV) and five from Section 17 of the Schedules for Clinical Assessment in Neuropsychiatry (SCAN) version 2.0. Clinicians rate the symptoms as either not present, suspected, definitely present, not relevant, or refused to answer. Study data were collected and managed using Research Electronic Data Capture (REDCap) hosted at the University of Bristol. REDCap is a secure, web-based software platform designed to support data capture for research studies (Harris et al. 2009). For our analyses of associations between GWC and psychotic experiences within at-risk groups, we included three PLIKS measures: 1) the total number of psychosis-like symptoms at age 24, 2) whether the young person ever had experienced none, suspected, or definite psychosis-like symptoms, including ‘definite non-clinical’ and ‘definite clinical’ diagnoses before age 24, and 3) whether the young person has none, suspected, or definite psychosis-like symptoms within the past 6 months at age 24, including ‘definite non-clinical’ and ‘definite clinical’ diagnoses. Please note that the study website contains details of all the data available through a fully searchable data dictionary and variable search tool (https://www.bristol.ac.uk/alspac/researchers/our-data/).

### 2.6 Statistical analysis

All statistical analyses were performed using the nonparametric statistical tool Permutation Analysis of Linear Models (PALM) in FSL (6.0.0) (Alberton et al. 2020). To correct for multiple comparisons, we used a false discovery rate (FDR) (Benjamini and Hochberg 1995) set at 5%. In the first set of analyses, we used general linear models to examine the effect of group on vertex-wise GWC, controlling for sex assigned at birth and age at the time of scan. Three separate analyses were performed to examine if there were group differences in GWC. First, we examined differences in GWC between the young adults who endorsed psychotic experiences and unaffected individuals in the PE study. Second, we examined if there were any differences in GWC between young adults in the high genetic-risk group and the low genetic-risk group in the SCZ-RbG study. Third, we examined if there were differences in GWC between the young adults who endorsed psychotic experiences and the genetic high-risk group, for this analysis excluding individuals who were part of both groups (n = 11).

In the second set of analyses, general linear models were used to examine whether the number (i.e., the total number of psychosis-like symptoms at age 24) and experiences of psychosis-like symptoms (i.e., in categories of whether the person ever had experienced none, suspected, or definite psychosis-like symptoms before age 24 and whether the person had none, suspected, or definite psychosis-like symptoms within the past 6 months at age 24) were associated with vertex-wise GWC within the young adults with psychotic experience and the genetic high-risk group, respectively, controlling for sex and age at scan. The number and experiences of psychosis-like symptoms variables were z-transformed prior to the analyses to improve numerical stability and interpretability.

## 3. Results

### 3.1 Demographics

The demographics and clinical characteristics of the samples are summarized in **Table 1**. The number of female participants was larger in both risk groups and the control groups. Participants in the psychotic experience group endorsed more psychotic experiences than participants in the high-genetic-risk group. Please see **Figure 1** for the distribution of psychotic experiences and age for all the groups.

**Figure 1.**
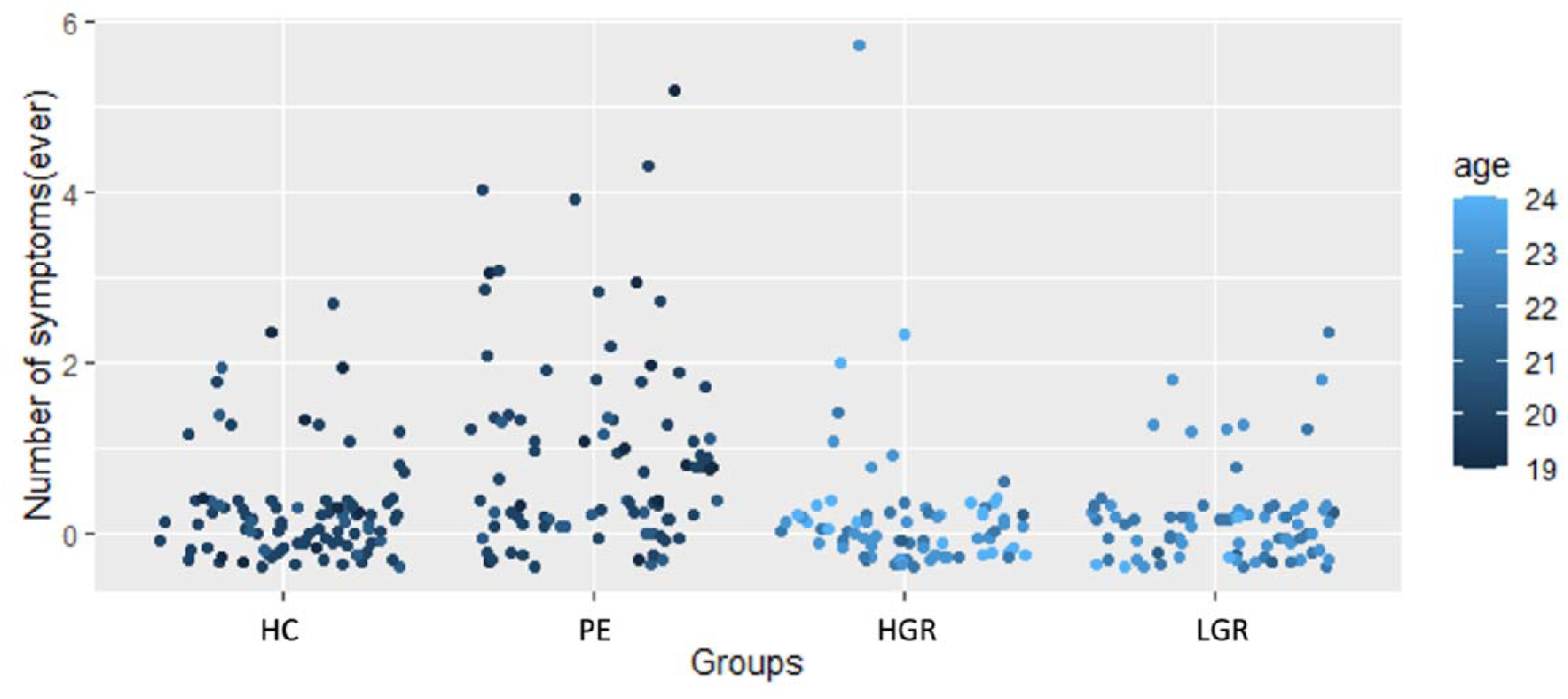
Distribution of the number of psychosis-like symptoms and age for the unaffected individuals (UI), psychotic experience group (PE), high genetic risk group (HGR), and low genetic risk group (LGR).

### 3.2 Group differences in grey/white matter contrast

In our first set of analyses, we used general linear models to examine group differences in vertex-wise GWC. No corrected significant group differences in GWC were observed between the psychotic experiences group and unaffected individuals or between the high genetic risk group and low genetic risk group. Moreover, no corrected significant differences in GWC were observed when comparing two risk groups, the psychotic experiences group and the high genetic risk group. Uncorrected effect sizes for all group comparisons are shown in **Figure 2**.

**Figure 2.**
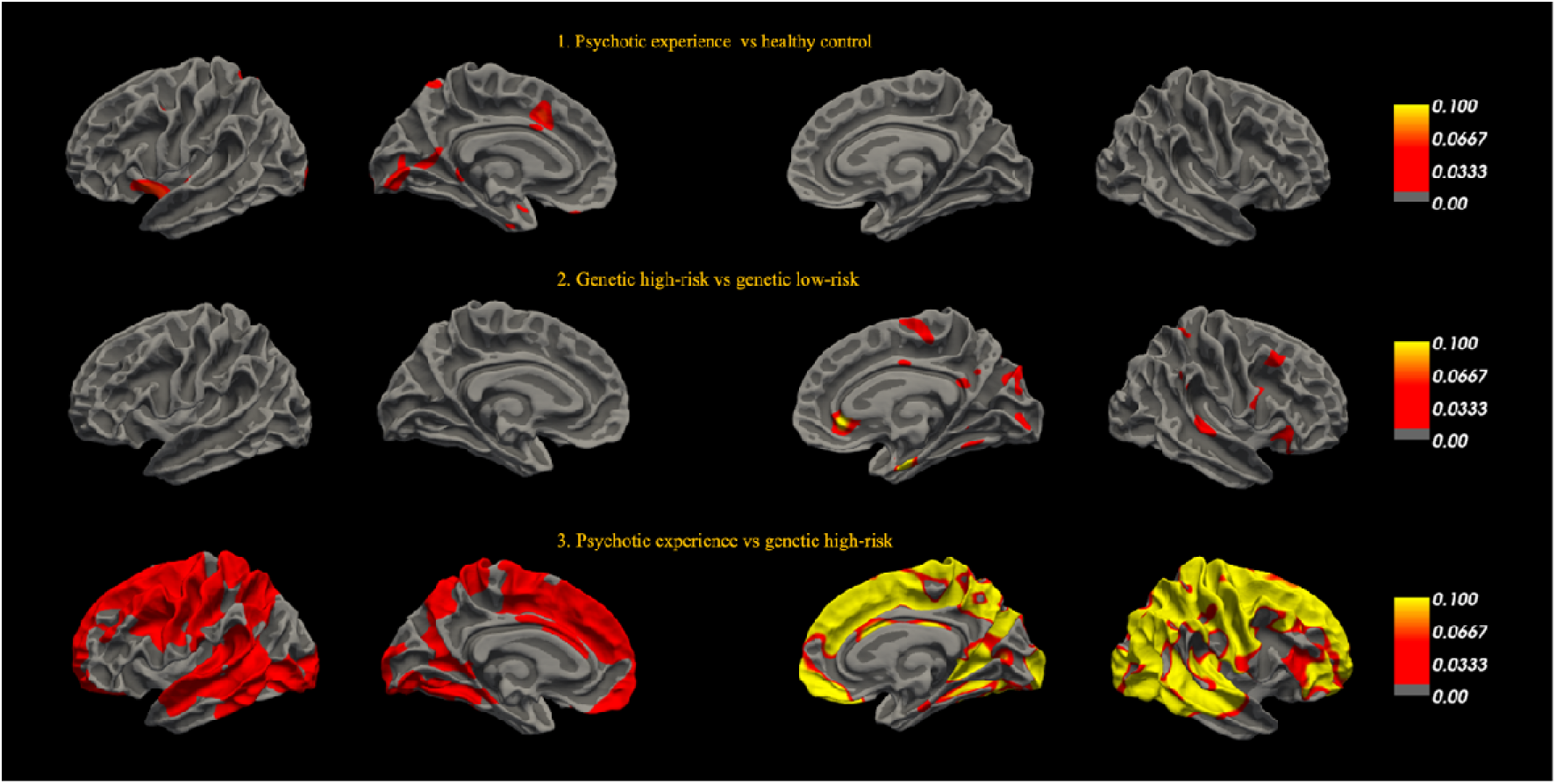
Uncorrected effect sizes for GWC group-level comparison. 1) Differences in GWC between individuals with psychotic experiences (PE) and unaffected individuals (HC), 2) Differences in GWC between individuals with high genetic risk (HGR) and low genetic risk (LGR) and 3) Differences in GWC between two risk groups, PE and HGR. Colour maps show the minimum and maximum thresholds.

### 3.3 Associations between gray/white matter contrast and psychotic-experiences

In our second set of analyses, we examined the association between vertex-wise GWC and the number of psychosis-like symptoms, experiences of psychosis-like symptoms before age 24, and experience of psychosis-like symptoms in the past six months within the two risk groups. The analyses did not show any corrected significant associations between the number or experience of psychosis-like symptoms and vertex-wise GWC in either of the two risk groups. Uncorrected effect sizes are shown in **Figure 3**.

**Figure 3.**
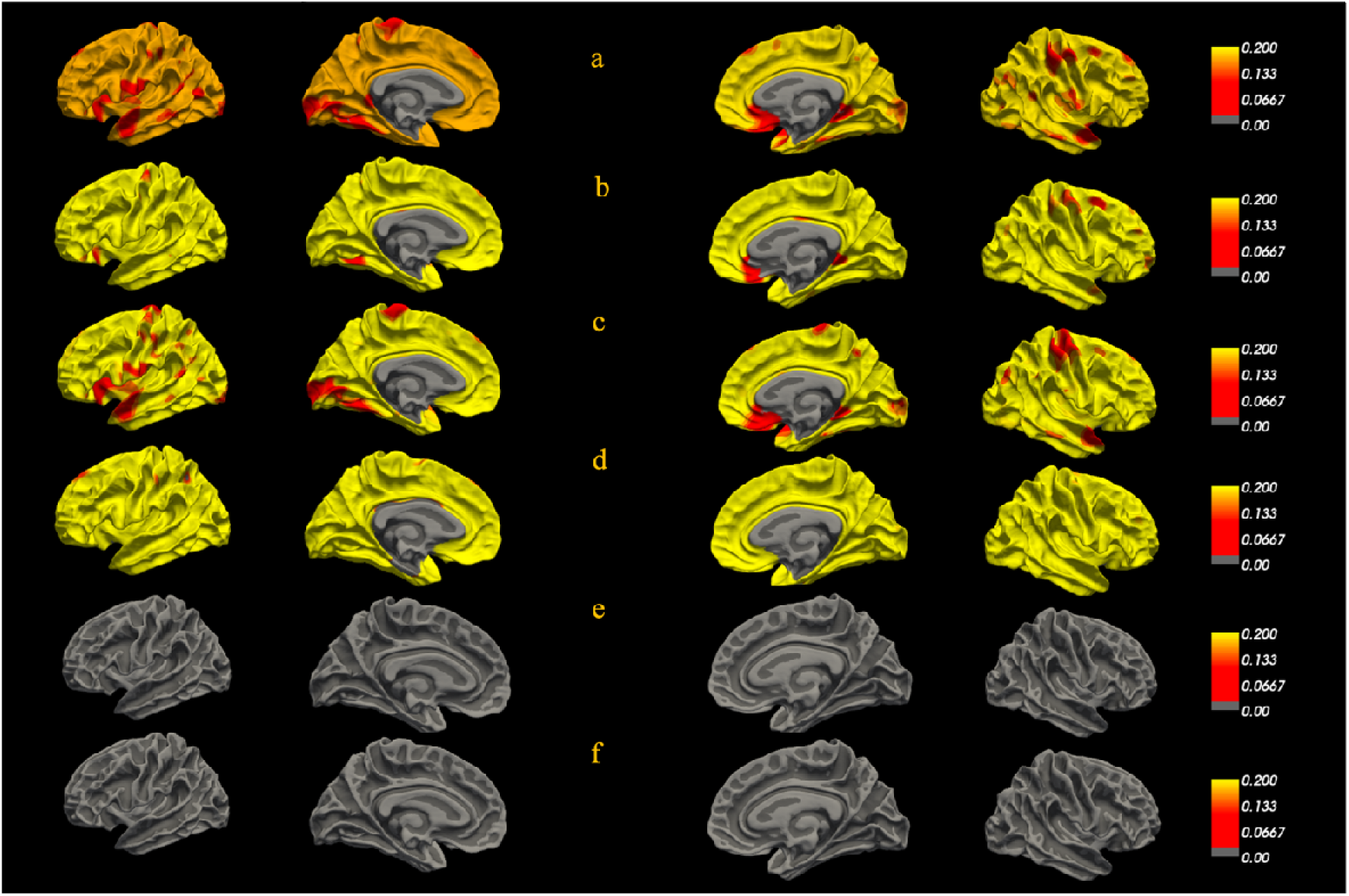
Uncorrected effect sizes for associations in the psychotic experience group between GWC and a) the number of psychosis-like symptoms, b) psychosis-like experiences within the past 6 months at age 24, and c) psychosis-like experiences before age 24. Associations in the genetic high-risk group between GWC and d) the number of psychosis-like symptoms, e) psychosis-like experiences within the past 6 months at age 24, and f) psychosis-like experiences before age 24. Colour maps show the minimum and maximum thresholds.

## 4. Discussion

The present study aimed to investigate brain microstructure as indexed by GWC in young adults who endorsed psychotic experiences or those at genetic risk for psychosis. Specifically, we tested for group differences between individuals who endorsed psychotic experiences and unaffected individuals, between individuals with high and low genetic risk, and between the two risk groups. No significant differences in GWC were observed between the groups. Our result, therefore, did not support our hypothesis that individuals with psychotic experiences would have regionally higher GWC in comparison to unaffected individuals and individuals with high genetic risk. Within the high-risk groups, individual differences in the number and experiences of psychosis-like symptoms were not significantly associated with GWC. Our hypothesis that psychotic experiences would be positively associated with GWC within a group endorsing psychotic experiences was thus not supported.

Previous studies conducted in clinical populations with psychotic disorders, specifically schizophrenia and bipolar disorder, have found group differences in GWC. Jørgensen et al. (2016) found higher GWC in pre- and postcentral gyri, the transverse temporal gyri, posterior insula, and parieto-occipital regions among patients with schizophrenia in comparison to unaffected individuals and patients with bipolar disorder. They also observed higher GWC in patients with bipolar disorder in comparison to unaffected individuals. In line with these results, Chwa et al. (2020) found higher GWC within the right superior frontal lobe encompassing the sensorimotor region in patients with schizophrenia in comparison to unaffected individuals. Kong et al. (2012; 2015), albeit with smaller samples, also observed differences in GWC among patients with schizophrenia and unaffected individuals, but in the opposite direction in large portions of the cortex, including frontal, temporoparietal and lateral occipital regions. We did not replicate these results in our sample of young adults with psychotic experiences or genetic risk for psychosis.

Although our study is the first one, to our knowledge, to use GWC to examine the underlying pathological process of psychosis in high-risk populations, other studies have examined brain structural (Chung et al. 2019; Del Re et al. 2021; Fornito et al. 2008; P. Fusar-Poli et al. 2011; Iwashiro et al. 2012; Klauser et al. 2015; Koutsouleris et al. 2009; Kwak et al. 2019; Mechelli et al. 2011; Sun et al. 2009; Takayanagi et al. 2017; Tomyshev et al. 2019; Velakoulis et al. 2006; Ziermans et al. 2009; Zikidi et al. 2020) and microstructural (Peters and Karlsgodt 2015; Smigielski et al. 2022; León-Ortiz et al. 2022; Drakesmith et al. 2016) differences in individuals at CHR for psychosis. Previous studies have associated the frequency and persistence of psychotic experiences with an increased risk of developing a psychotic disorder (Kaymaz et al. 2012). However, there is an ongoing debate about whether these experiences and the brain changes observed in young adults with heightened risk for psychosis are markers of early neurodevelopmental insults and can be considered vulnerability factors or whether they occur later, coinciding with the manifestation of PEs, symptom deterioration, or medication use (Fonville et al. 2019; P. Fusar-Poli et al. 2011). Moreover, potential sampling bias in studies of at-risk individuals must be considered (Jalbrzikowski et al. 2021; Smieskova et al. 2010b). Help-seeking individuals with CHR have a 15% risk of developing psychosis within 38 months, compared to a 0.1% risk in the general population during the same period (Paolo Fusar-Poli et al. 2016). There is a possibility that pathological processes are more advanced in help-seeking individuals with prodromal symptoms, especially among those who later convert to psychosis (Cannon 2008). In line with this, studies have shown that abnormalities in the subcortical white matter among patients with schizophrenia developed as the disorder progressed and were associated with a longer duration of illness (Bartzokis, 2011; Friedman et al., 2008). Future research should examine GWC development in CHR populations and compare converters and non-converters as the maturation of myelination in adolescents overlaps with the period of developmental vulnerability of psychotic disorders (Chafee and Averbeck 2022).

Similar challenges arise within clinical populations, as most neuroimaging studies have been focused on chronically ill, medicated patients with schizophrenia and bipolar disorder (Birur et al. 2017). This complicates the differentiation of vulnerability characteristics from those influenced by disease progression or the effects of psychotropic medications (Birur et al. 2017). Medication-induced metabolic syndromes can lead to microinfarctions and disordered perfusion in the subcortical white matter (Mighdoll et al. 2015). Jørgensen et al. (2016) found a positive association between GWC and experiences of hallucinations in the transverse temporal cortices bilaterally and subthreshold symptoms in the transverse temporal cortex and left occipital lobe clusters. Chwa et al. (2020) found a negative association between cumulative dosage exposure to second-generation antipsychotics and GWC bilaterally in the frontal lobe and within the right orbital frontal cortex after controlling for cortical volume differences. These observations open avenues for explaining the GWC differences observed in clinical populations are related to secondary factors such as the severity of clinical symptoms or medication. Previous studies in both clinical and high-risk populations have reported associations between the severity of symptoms and brain structure and microstructure (Satterthwaite et al. 2016; Drakesmith et al., 2016).

GWC appears to be a marker for both early development and later age-related decline and might capture pathological processes in both younger and older ages. Studies have shown that GWC predicts age accurately in youth, reinforcing its role in tracking early brain development (Lewis, Evans, and Tohka 2018; Norbom et al. 2019). GWC has also been found to capture individual differences linked to prodromal psychosis and cognitive ability (Norbom et al. 2019) and differences in children with attention deficit hyperactivity disorder (Wang et al. 2023), indicating its potential to identify early neurodevelopmental alterations.

On the other hand, increased age is also associated with a decline in GWC (Salat et al. 2009; Vidal□Piñeiro et al. 2016). In older clinical populations, GWC has been proven to be sensitive to capturing β-amyloid pathology in earlier stages of Alzheimer’s disorder (AD) (Putcha et al. 2023). GWC is suggested to capture microstructural changes before the emergence of cortical atrophy and may be more sensitive to early microstructural changes than conventional neuroimaging parameters (Putcha et al. 2023; Xu et al. 2024). Further, changes in GWC have been associated with cognitive performance among AD individuals with underlying β-amyloid pathology and with increased age among individuals with dementia (Salat et al. 2009; Xu et al. 2024). These findings indicate that GWC may serve as a marker for both early developmental changes and later age-related decline, with potential clinical utility in capturing shifts across the lifespan.

Our results are in line with our hypothesis that there would not be any differences in GWC between individuals with high and low genetic risk for psychosis. Results from previous studies examining the association between schizophrenia PRS and brain structure and microstructure are contradictory (Dimitriadis et al. 2023; Fonville et al. 2019; Lancaster et al. 2019; Neilson et al. 2018; Ohi et al. 2014). A meta-analysis did not find any association between schizophrenia PRS and brain structure, and studies that found an association reported small effect sizes (van der Merwe et al. 2019). This could be due to the limited predictivity power of schizophrenia PRS. PRS performs worse when the sample consists of individuals from different ancestries (Andreassen et al. 2023), however, our sample consisted mainly of white European individuals. A previous ALSPAC study did not find any association between schizophrenia PRS and psychotic experiences (Jones et al. 2016), while other studies with larger samples have found associations between schizophrenia PRS and psychotic-like experiences (Barbu et al. 2023; Elkrief et al. 2023).

The null results of the present study should be interpreted in the light of some limitations. As discussed above, our samples endorsed relatively few psychotic experiences and should, therefore, be considered as a low-risk group. Second, larger samples and greater statistical power might be needed to detect subtle differences in GWC clinical or genetic risk groups. Third, we did not investigate the differences in GWC between individuals who transitioned to full psychotic disorder and those who did not. Fourth, the recruitment of individuals with psychotic experiences from the general population has lower predictive power than the recruitment of help-seeking participants, which can lead to null results (Paolo Fusar-Poli et al. 2015). However, population-based sampling is argued to minimise self-reporting, referral and selection biases (Drakesmith et al. 2015). Fifth, an older GWAS (Ripke et al. 2014) was used to calculate the polygenic risk score for schizophrenia, which could limit the predictive power of our study. Lastly, we did not control for medication and drug use. Studies have shown that substance use have a negative effect on connectivity between brain regions and altered white matter structure (Malone, Hill, and Rubino 2010; S. Y. Chan et al. 2022). Considering these limitations, future studies could focus on the developmental nature of psychosis using a longitudinal study design including different imaging metrics and clinical measures at multiple time points and seek to include larger samples that also include individuals with higher clinical or genetic risk.

## 5. Conclusion

The findings of the current study showed no GWC differences between young adults with psychotic experiences and unaffected individuals or between young adults with high and low genetic risk for psychosis, nor any association between GWC and the number or experiences of psychosis-like symptoms. This suggests that detecting differences in brain microstructure in individuals with relatively low psychosis risk is challenging using GWC.

## Data Availability

All data are available from the Avon longitudinal study of parents and children upon request.

## Acknowledgements

We are extremely grateful to all the families who took part in this study, the midwives for their help in recruiting them, and the whole ALSPAC team, which includes interviewers, computer and laboratory technicians, clerical workers, research scientists, volunteers, managers, receptionists and nurses.

